# ARDSFlag: An NLP/Machine Learning Algorithm to Visualize and Detect High-Probability ARDS Admissions Independent of Provider Recognition and Billing Codes

**DOI:** 10.1101/2022.09.27.22280416

**Authors:** Amir Gandomi, Phil Wu, Daniel R Clement, Jinyan Xing, Rachel Aviv, Matthew Federbush, Zhiyong Yuan, Yajun Jing, Guangyao Wei, Negin Hajizadeh

## Abstract

Acute respiratory distress syndrome (ARDS) is a type of respiratory failure characterized by bilateral pulmonary infiltrates that cannot be explained entirely by cardiogenic pulmonary edema. ARDS is the primary cause of mortality in COVID-19 patients and one of the leading causes of morbidity and mortality in ICUs. Despite its significance and prevalence, the detection of ARDS remains highly variable and inconsistent. In this work, we develop a tool to automate the diagnosis of ARDS based on the Berlin definition to increase the accuracy of ARDS detection using electronic health record (EHR) fields. ARDSFlag applies machine learning (ML) and natural language processing (NLP) techniques to evaluate Berlin criteria by incorporating structured and unstructured data. The output is the ARDS diagnosis, onset time, and severity. We have also developed a visualization that helps clinicians efficiently assess ARDS criteria retrospectively and in real time. The method includes separate text classifiers trained using large training sets to find evidence of bilateral infiltrates in radiology reports (accuracy of 91.9%±0.5%) and heart failure/fluid overload in radiology reports (accuracy 86.1%±0.5%) and echocardiogram notes (accuracy 98.4%±0.3%). A holdout set of 300 cases, which was blindly and independently labeled for ARDS by two groups of clinicians, shows that the algorithm generates an overall accuracy of 89.0%, with a specificity of 91.7%, recall of 80.3%, and precision of 75.0%. Compared with two other ARDS identification methods used in the literature, ARDSFlag shows higher performance in all accuracy measures (an increase of 25.5% in overall accuracy, 6.5% in specificity, 44.2% in recall, 31.7% in precision, and 38.20% in *F*_1_-score over the best of the two detection methods).

## 1. Introduction

Acute respiratory distress syndrome (ARDS) is a syndromic disease of inflammatory lung injury without a sensitive and specific diagnostic test. ARDS is associated with a high mortality rate (∼40%) and substantially impacts survivors’ quality of life ^1,2^. The definition of ARDS has evolved from the original definition in 1967 to the more recent 2012 Berlin criteria ^3,4^. The diagnosis of ARDS based on the Berlin definition requires a constellation of clinical findings, including the multi-lobar involvement of the lungs and the origin of parenchymal changes and hypoxemia. As a result, variability in the detection of ARDS remains problematic both in clinical practice and research^5–9^.

For example, the LUNG SAFE study, which was the largest multicenter cohort study of ARDS patients to investigate the epidemiology and outcomes of ARDS across 459 ICUs from 50 countries ^6^, found that, on average, 40% of ARDS patients identified by an automated algorithm using the Berlin criteria were not diagnosed by the clinicians. In addition, there was a delay in diagnosing ARDS among 66% of patients ^6^. Early diagnosis of ARDS enables timely implementation of protective lung ventilation strategies and adjunctive measures ^10^, leading to lower mortality rates ^6,11,12^. Furthermore, consistency in ARDS detection enables investigators to study the associations of treatment trajectories and patient characteristics with outcomes ^13^.

This study contributes to the ARDS literature by developing *ARDSFlag*, a new method to automate the detection of ARDS based on structured and unstructured textual data stored in electronic health record (EHR) systems. ARDSFlag uses machine learning (ML) and natural language processing (NLP) techniques to evaluate Berlin criteria. ML and NLP have been proven to offer strong potential for identifying and predicting complex medical conditions by incorporating EHR data ^14–17^. We also develop a visualization that integrates all components of the Berlin criteria in one graph. The use of this visualization may enhance the efficiency and accuracy of clinicians in detecting ARDS cases.

ARDSFlag evaluates the four parameters of the Berlin definition. It includes separate text classifiers trained using large training sets to detect bilateral infiltrates (BI) in radiology reports and heart failure/fluid overload (HF/FO) in radiology and echocardiogram (echo) reports. We use a validation set of 100 cases, developed by an independent review of two groups of clinicians, to find the optimal temporal sequence of Berlin parameters. Using a separate ground truth set of 300 cases, we show that the algorithm outperforms other methods in the literature. The competing methods include the use of International Classification of Diseases (ICD) codes and the method developed by Serpa Neto et al.^18^

## 2. Methods

### 2.1. Dataset

We used the Medical Information Mart for Intensive Care III (MIMIC-III) dataset ^19^ to develop and test the automated ARDS detection algorithm. We used hospital admissions as the unit of analysis and, as shown in Figure 1, limited the cohort to adult admissions (age≥ 18). Since the Berlin definition is based on chest imaging reports, partial pressure of arterial oxygen (*PaO*_2_), fractional inspired oxygen (*FiO*_2_), and positive end expiratory pressure (*PEEP*), the cohort is further limited to admissions with at least one record of each. The inclusion criteria led to an initial cohort of 19,534 admissions.

**Figure 1.**
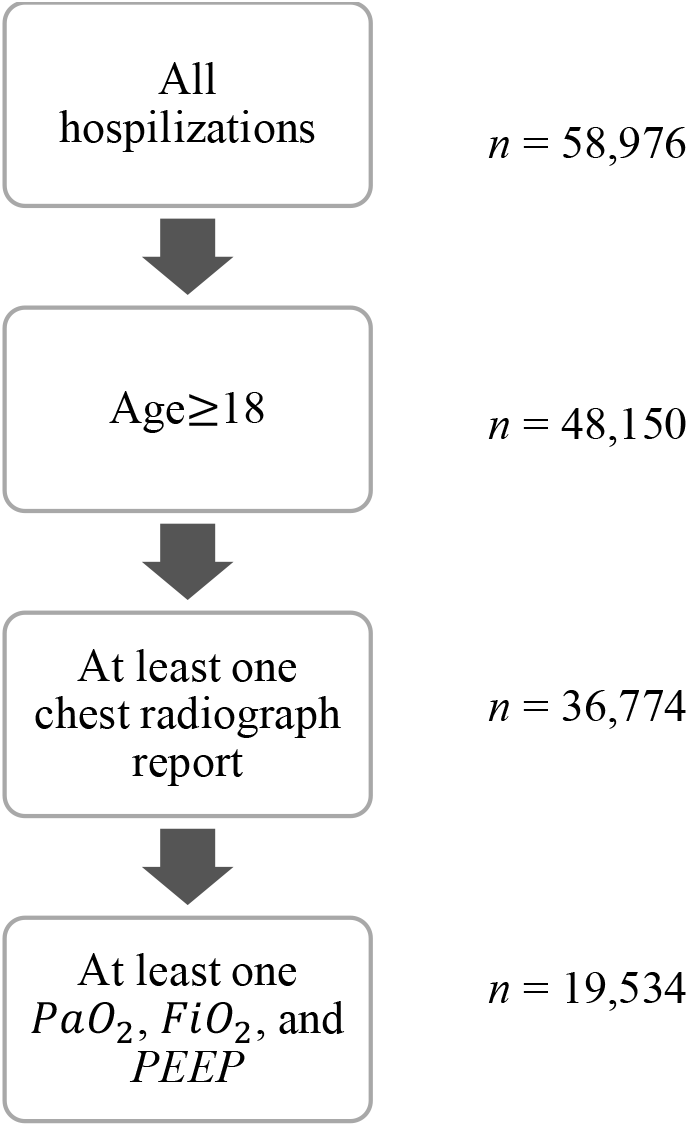
Cohort selection

Each patient’s relevant data points were fetched from MIMIC’s different tables and stored in individual multi-dimensional time series. We conducted extensive querying to find the correct items associated with each variable and to cross-validate the data in different tables. Since MIMIC is sourced from two different clinical information systems (MetaVision and CareVue), validation had to be completed twice. When possible, we externally validated the target data. For instance, we used LOINC codes to verify lab items.

A column in the time series data captured the ICU stay’s significant events, including admission, intubation, extubation, discharge, expiration, and disposition. We used Makhnevich et al.’s ^20^ algorithm to find the accurate intubation time. Another algorithm was developed to find the time of extubation based on recorded procedures, ventilator parameters, and oxygen delivery methods. Dispositions were mapped into four general categories *expired, hospice, home*, and *facility*, where the latter refers to locations such as skilled nursing facility, rehab, and short-/long-term hospital.

Central to the detection of ARDS is the evidence of bilateral infiltrates (BI) in chest radiographs and the diagnosis of cardiac failure or fluid overload (*HF*/*FO*). The following section explains that we have developed NLP algorithms to extract such evidence from patient notes. Positive incidences of BI and *HF*/*FO* are also stored in the time series’ Event column.

The data preprocessing pipeline was developed using Python 3.6. All factors related to ARDS detection are visualized in a single graph referred to as the *ARDS graph* hereafter. A sample ARDS graph is presented in Figure S1. The corresponding time series is available in CSV format in supplementary files.

To increase the efficiency of manual chart reviews, we developed the pipeline to print the structured admission data (e.g., demographics, admission and discharge dates, and the initial diagnosis), the ARDS graph, and all relevant notes for every case in a single PDF file. A set of keywords were selected to be highlighted in the pdf file. This data presentation method significantly increased the efficiency of manual chart reviews to evaluate the algorithm’s accuracy.

### 2.2. ARDS detection algorithm

#### 2.2.1. Overview of the algorithm

Based on the Berlin criteria ^21^, ARDS is defined by: (1) acute onset, (2) *P*/*F* ≤ 300 *mm Hg* while *PEEP* ≥ 5 *cm H*_2_*O*, (3) BI in chest radiographs, and (4) the absence of HF/FO as the primary origin of pulmonary edema.

Following Serpa Neto et al.^18^ and Le et al.^8^, we used tracheostomy as a proxy to evaluate the first condition: if a patient had a tracheostomy within seven days of admission, they were classified as non-ARDS. Furthermore, the time of ARDS onset (determined based on the second and third criteria) must be within seven days after the first record of receiving *PEEP* ≥ 5.

For the second condition, we used the time series data. *PaO*_2_ measurement times do not always coincide with *FiO*_2_ or *PEEP* reporting times in the patient chart. In such cases, we carried forward the last reported *FiO*_2_ and *PEEP* level to calculate *P*/*F* ratio. This procedure assumes that all changes in the ventilator parameters are recorded in the patient chart.

We trained different text classifiers to evaluate the third and fourth conditions. A text classifier was trained to find evidence of BI in chest radiology reports. Separate classifiers were developed to find *HF*/*FO* in chest radiology and echo reports. Text classifiers are detailed in the next section.

Figure 2 depicts the logic for the sequence of conditions. As shown in the top graph in Figure 2a, evidence of BI is generally valid within *T*_*BI*_ ± *δ*_*BI*_, where *T*_*BI*_ is the time of the radiology and *δ*_*BI*_ is the BI time window. A low *PP*/*FF* ratio counts toward ARDS diagnosis if it occurs within this boundary. If there is another chest radiology *without* evidence of BI within *T*_*BI*_ ± *δ*_*BI*_, as shown in the two bottom graphs in Figure 2a, the boundary shrinks. As discussed later on, the optimal value of *δ*_*BI*_ is found to be one day.

**Figure 2.**
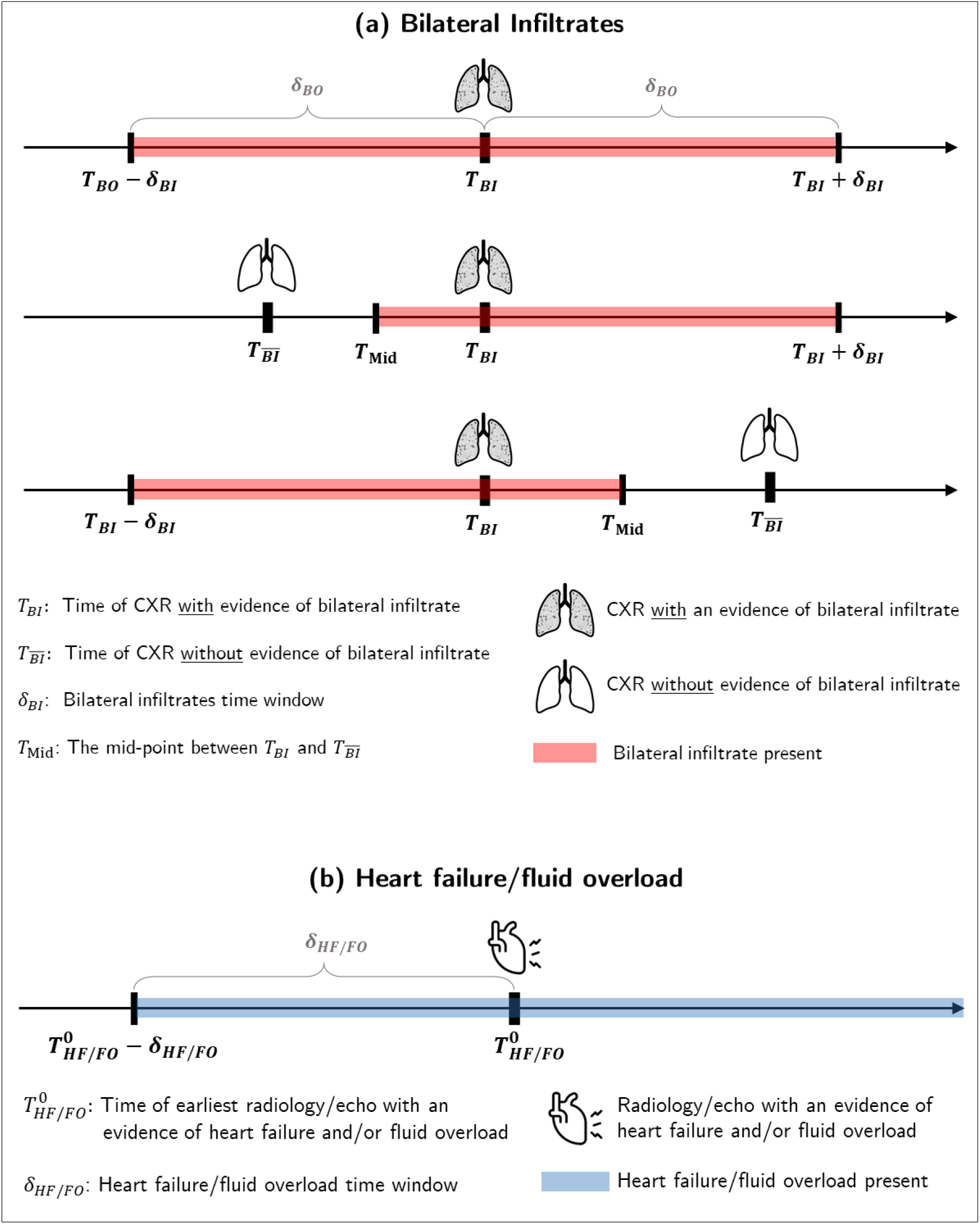
The timing of ARDS conditions: a. BI, b. Heart failure/fluid overload

Figure 2b shows the logic for the origin of edema. Let 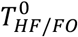 denote the time of the earliest echo/CXR with evidence of HF/FO, *δ*_*HF*/*FO*_ denote the corresponding time window, and *T*_0_ denote the time of the potential ARDS onset (the earliest time the first three ARDS conditions are satisfied). If 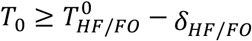 then HF/FO is identified as the origin of respiratory failure. Otherwise, *T*_0_ is the time of ARDS onset. The optimal value of *δ*_*BI*_ will be shown to be five days. Figure S2 further explains the Berlin implementation logic using a few sample cases.

To reduce the false-positive rate, we included the length of mechanical ventilation as an additional criterion; patients who received less than 48h of mechanical ventilation were excluded unless they expired or were discharged to hospice within 48h after intubation or extubation. Thus, we do *not* exclude short ventilations that are a result of severe illness (expire within 48h after intubation) or elective extubation (expire or discharge to hospice within 48h after extubation). Serpa Neto et al.^18^ exclude patients receiving less than 48h of ventilation regardless of whether the ventilation terminated due to death or elective palliative care, which may lead to the omission of severe cases. We perform a sensitivity analysis on this condition in the Discussion section.

In what follows, we describe the classifiers built to detect BI and *HF*/*FO*.

#### 2.2.2. Detection of Bilateral infiltrates (BI)

We trained a sentence-based classifier to detect the evidence of BI in chest radiograph reports. A report is classified as positive if it includes at least one positive sentence. Figure S3 shows the process of developing the training set, which included 2,376 sentences.

Two clinicians labeled all the sentences independently as “positive” (i.e., providing evidence of BI) or “negative.” “Positive” was defined as including mention of both right and left lungs involvement of the following: infiltrate, opacity, consolidation, airspace disease, aspiration, and pneumonia. Unilateral lung involvement, presence of bilateral pleural effusion, and consolidations attributed directly to atelectasis only were labeled negative. If the radiologist qualified an improvement or worsening of BI, the sentence was labeled as positive. Conversely, the sentence was labeled negative if the impression qualified interval resolution or recovery of BI.

Clinician’s agreement rate was 88.0%. Inconsistencies were resolved by deliberation with other clinicians in the group. Furthermore, the group consulted a diagnostic radiologist to provide insight into the decision-making. Finally, 938 positive sentences (positive rate = 39.5%) were identified in the training set. Figure S4 shows a summary of the training set. The data were divided into train and test sets using stratified sampling at a 75:25 ratio.

We built a classification pipeline with three main steps: text preparation, vectorization, and classification. For each step, we experimented with different parameter settings and used grid search with five-fold cross-validation to find the architecture that returns the maximum *F*_1_ score for the positive class. Table S1 lists all pipeline parameters.

The text preparation step involves removing tags, punctuations (except for question marks), numbers, and multiple whitespaces, unifying all variations of common phrases into a single form (e.g., ‘*please’* and ‘*pls’*, ‘*pneumonia’* and ‘*pna’*, ‘*campared to’* and *‘in comparison with’*), and converting common multi-word phrases into unigrams (e.g., *‘pulmonary edema’* to’ *pulmonaryedema’, ‘consistent with’* to ‘*consistentwith’*, and, ‘*final report*’ to *‘finalreport’*) and replacing the results of MIMIC’s named entities with a generic name (e.g., replacing *[**Doctor Last Name 107**]* with *LastName*). We varied two parameters in this step: using Standard English versus a customized list of stopwords and whether to apply stemming or not.

We tested two approaches for vectorization, bag of words (BoW) and word embedding. For BoW, we implemented the term frequency-inverse document frequency (TF-IDF) weighting scheme. We experimented with TF-IDF parameters listed in Table S1. The word embedding was implemented using *spaCy*’s pre-trained word vectors. The vector representation of each sentence was obtained by averaging its token vectors. We used singular value decomposition (SVD) for dimensionality reduction and incorporated the number of dimensions as a parameter in the grid search.

We examined six learning models for classification and experimented with their hyperparameters, summarized in Table S1. It is worth noting that Stochastic Gradient Descent (SGD) is an optimization technique for the training of different linear classifiers rather than a learning model by itself ^22^. For instance, SGD with the hinge loss function is equivalent to a linear support vector machine (SVM).

The grid search within the pipeline parameters’ space returned TF-IDF vectorization and SGD with the modified Huber loss function as the optimal configuration. Details of the optimal setting are highlighted in Table S1. We replicated the optimal classification pipeline 30 times with different random seeds for test/train split and shuffled the training after each epoch. Table 1 shows the summary for different accuracy metrics. Accuracy is measured based on the 25% of the data set aside for testing in each replication.

**Table 1.** Accuracy of the BI classifier (based on 30 random test/train splits of 2,376 sentences)

| Class | Measure | Min | Max | 95% Confidence Interval |
| --- | --- | --- | --- | --- |
| Overall | Accuracy | 89.1% | 94.1% | 91.9% ± 0.5% |
| Negative | Precision | 90.8% | 97.6% | 94.9% ± 0.5% |
| | Recall | 88.3% | 95.5% | 91.6% ± 0.7% |
| | $F_1$ | 91.0% | 95.1% | 93.2% ± 0.4% |
| Positive | Precision | 84.2% | 93.0% | 87.8% ± 0.8% |
| | Recall | 86.0% | 96.6% | 92.4% ± 0.8% |
| | $F_1$ | 86.1% | 92.6% | 90.0% ± 0.6% |

#### 2.2.3. Detection of Heart failure/fluid overload (HF/FO)

Following a similar approach, we developed a pipeline to extract evidence of HF/FO in chest radiology and echo reports. Echo reports tend to have a different syntax and lexicon than radiology reports. Hence, we developed separate classifiers for radiology and echo reports. The keywords used to find the relevant sentences are *cardiac shock, cardiac arrest, cardiac failure, fluid overload, volume overload, heart failure, CHF, hydrostatic, cardiogenic, hypervolemia, systolic dysfunction, diastolic dysfunction, LVSD*, and *LVDD*. After reviewing an initial training set, we decided to include the sentences before and after the focal sentence to capture the context better. Thus, the HF/FO classifier’s input is a three-sentence document where the keywords occur in the middle unless the keyword is in the first or last sentence, resulting in a two-sentence document.

For the radiology reports classifier, a training set of 2,000 documents was randomly generated from the patients’ study cohort. To achieve a representative variety, we picked a maximum of two documents per patient. Two clinicians labeled the documents blindly. CHF positivity was defined by the phrase or combination of phrases that suggested a cardiogenic etiology or stated the presence of heart failure, congestive heart failure (CHF), edema, or fluid or effusions. Reports that did not comment on pulmonary parenchyma or collectively did not suggest a cardiogenic etiology responsible for pulmonary abnormalities were labeled negative. The disagreements, which had a rate of 6.2%, were settled by discussion and consensus. The final training set included 1,808 documents with 1,020 positive examples (positive rate = 56.4%). We stratified-split the data into train and test at an 80:20 ratio. Figure S5 summarizes the HF/FO classifier’s training set.

Following a similar procedure, we developed a training set for the echo reports classifier, which consisted of 1,048 random documents with 534 positive examples (positive rate = 50.1%).

Similar to the BI pipeline, we conducted an extensive grid search with five-fold cross-validation to find the optimal pipeline structure for the two HF/FO classifiers. Table 2 summarizes the test data accuracy levels obtained in 30 replications of the grid search.

**Table 2.** Accuracy of the HF/FO classifiers (based on 30 random test/train splits of 2,000 radiology and 1,048 echo documents)

| Data source | Class | Measure | Min | Max | 95% Confidence Interval |
| --- | --- | --- | --- | --- | --- |
| Radiology reports | Overall | Accuracy | 83.4% | 88.7% | 86.1%±0.5% |
|  | Negative | Precision | 80.5% | 89.7% | 85.1%±0.9% |
|  |  | Recall | 77.7% | 88.3% | 82.5%±0.9% |
| | | $F_1$ | 81.1% | 86.6% | 83.8%±0.6% |
|  | Positive | Precision | 84.3% | 90.6% | 86.8%±0.6% |
|  |  | Recall | 84.7% | 92.5% | 88.8%±0.8% |
| | | $F_1$ | 85.2% | 90.2% | 87.8%±0.5% |
| Echo reports | Overall | Accuracy | 96.6% | 99.6% | 98.4%±0.3% |
|  | Negative | Precision | 96.1% | 100.0% | 98.7%±0.4% |
|  |  | Recall | 95.3% | 100.0% | 98.0%±0.4% |
| | | $F_1$ | 96.5% | 99.6% | 98.3%±0.3% |
|  | Positive | Precision | 95.7% | 100.0% | 98.1%±0.4% |
|  |  | Recall | 96.3% | 100.0% | 98.7%±0.4% |
| | | $F_1$ | 96.6% | 99.6% | 98.4%±0.3% |

#### 2.2.4. Parameter tuning

We used a random set of 400 admissions to tune the algorithms’ parameters and evaluate their accuracy. In order to address the class imbalance issue, 100 of the 400 cases were randomly selected from a cohort that was classified as positive for ARDS by an initial version of the algorithm. The tuning parameters are *δ*_*BI*_ and *δ*_*HF*/*FO*_, the BI and HF/FO time windows. Two groups of clinicians were instructed to follow Berlin criteria and independently label cases for ARDS objectively. Each case’s relevant data was presented in a PDF file as described in Section 3. Disagreements were settled by a joint evaluation (rate = 9%). We used 100 cases (25%) for parameter tuning and the remaining 300 cases as a holdout set to estimate the algorithm’s accuracy.

For the parameter tuning, we performed a grid search over values of *δ*_*BO*_ and *δ*_*HF*/*FO*_ in the set {4h, 8h, 12h, 1d, 2d, 5d, 7d} and used *F*_1_-score to find the best combination of parameter values. 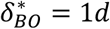 and 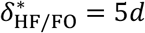 generated the best result with *F*_1_ = 84% (Accuracy = 92%, Precision = 80.8%, Recall = 87.5%).

The optimal time windows (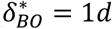 and 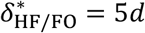) are clinically relevant. ARDS is characterized by rapid onset of bilateral infiltrates that can take weeks to months to fully resolve in most cases. Patients with fluid overload may initially meet Berlin criteria; however, the bilateral infiltrates seen will usually improve rapidly with medical management. If a resolution of bilateral infiltrates occurs within a matter of days, it is likely to result from a cardiogenic process or fluid overload as opposed to ARDS. Table 3 shows the summary of the grid search results, which is based on 49 pairs of *δ*_*BO*_ and *δ*_*HF*/*FO*_ values.

**Table 3.** Results of the grid search to find optimal time windows (*δ*_*BO*_and *δHF*/*FO*) for ARDS detection algorithm

| Measure | Min | Max | 95% Confidence Interval |
| --- | --- | --- | --- |
| Accuracy | 78.0%, | 92.0% | 84.4%±1.3% |
| Precision | 62.5%, | 91.7% | 84.5%±2.2% |
| Recall | 77.6% | 93.4% | 84.3%±1.8% |
| $F_1$ | 53.1%, | 80.8% | 64.6%±2.9% |

## 3. Results

Table 4 presents the confusion matrix for ARDSFlag based on the 300 holdout cases in the test set. The overall accuracy of the algorithm is 89%. There were 71 true positive cases (defined based on the manual review by two groups of clinicians), 57 of which were detected by the algorithm (recall = 80.3%). The precision for the positive class is 75% leading to an *F*_1_ score of 77.6%. There were 229 true negative cases, 210 of which were correctly classified (specificity = 91.7%).

**Table 4.** Confusion matrix for the holdout set. The predicted label is shown in rows. Columns show the true label determined based on an independent review of two groups of clinicians (e.g., the algorithm generated 57 true positive and 14 false negative cases)

| Predicted Label | True Label |  | Total |
| --- | --- | --- | --- |
|  | Positive ARDS | Negative ARDS |  |
| Positive ARDS | 57 | 19 | 76 |
| Negative ARDS | 14 | 210 | 224 |
| Total | 71 | 229 | 300 |

Table 5 shows the algorithm performance compared to the two other methods used in the literature: automated implementation of the Berlin criteria developed by Serpa Neto et al.^18^ and the use of International Classification of Diseases (ICD) codes to define ARDS^23–26^. We reproduced the Serpa Neto et al.^18^’s method using its detailed description in Le et al.^8^ and confirmed the results match. For ICD-based algorithms, we included patients with respiratory failure as their primary or secondary diagnosis (ICD-9 codes 518.51, 518.52, 518.81, and 518.82) and experimented with the inclusion of mechanical ventilation procedure (ICD-9 codes 96.70, 96.71, 96.72) (similar to, e.g., Schwager et al.^23^ and Eworuke et al.^24^), and exclusion of patients with a primary diagnosis of heart failure (ICD-9 codes 410, 411, 412, 414, 428) (similar to, e.g., Liu et al.^27^ and TenHoor et al., 2001^26^). The highest accuracy was achieved by incorporating the respiratory failure and heart failure codes and not including the ventilation procedure. Table 5 shows the outcome of this optimal ICD configuration. The results show that the algorithm outperforms other methods in all measures.

**Table 5.** Comparison of accuracy of different ARDS detection methods

| Method | Accuracy | Specificity | Recall | Precision | $F_1$-score |
| --- | --- | --- | --- | --- | --- |
| Proposed algorithm | 89.0% | 91.7% | 80.3% | 75.0% | 77.6% |
| ICD-9 | 60.0% | 68.1% | 33.8% | 24.7% | 28.6% |
| Serpa Neto et al.<sup>18</sup> | 73.5% | 85.2% | 36.1% | 43.3% | 39.4% |

The extent of overlap among the three methods is depicted using Venn diagrams in Figure 3. Figure 3a shows their relationship over the 71 true positive cases in the test set. Our proposed algorithm (ARDSFlag) detected 57 (80.3%) ARDS cases, 27 (38.0%) of which are missed by both ICD-9 and Serpa Neto et al.^18^ methods. However, ARDSFlag failed to detect 8 (11.3%) true positive cases that were identified by either one of the other two methods. All three methods failed to detect 6 (8.4%) true ARDS cases as defined by manual review by two groups of clinicians. Figure 3b shows the overlap among positive cases identified by the three methods within the 300 admissions in the test set, regardless of their true label. Collectively, the three methods detected 186 positive cases with an agreement rate of 4.8% (*n* = 9).

**Figure 3.**
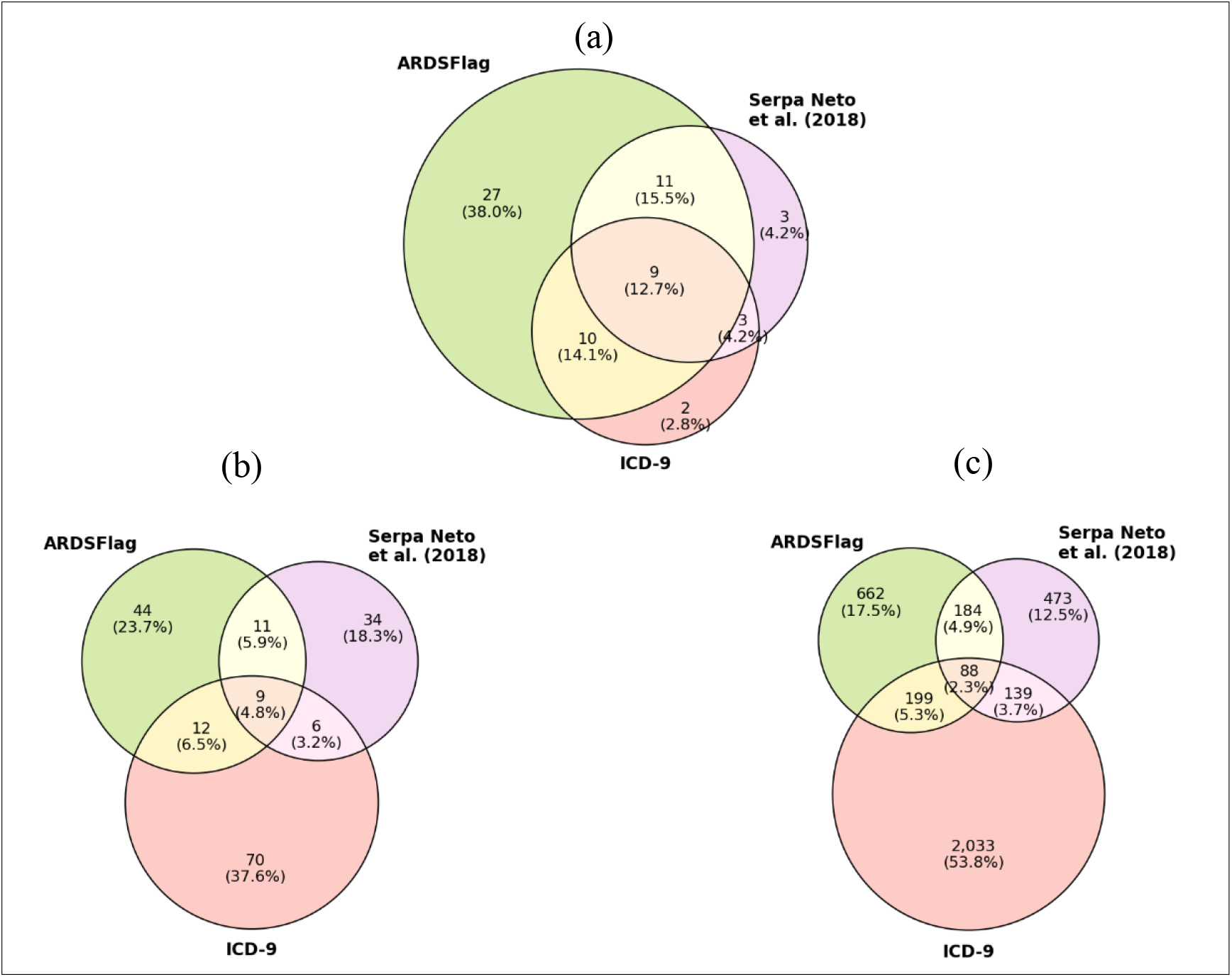
Venn diagram of positive ARDS cases detected by three methods within **a**. the true positive cases in the test set (*n* = 71), **b**. all holdout cases (*n* = 300), and **c**. the entire cohort (*n* = 19,534). The

As evident from Figure 3b, ICD-9 over-detects ARDS cases and Serpa Neto et al.^18^ under-detects ARDS. We used the three methods to find ARDS cases in the entire study cohort and evaluate whether this pattern is generalizable. Figure 3c shows the results for the entire study cohort (*n* = 19,534). The ARDSFlag detected a total of 1,133 ARDS cases (incidence rate of 5.8%), ICD-9 resulted in 2,459 (rate = 12.6%), and Serpa Neto et al.^18^ generated 884 (rate = 4.5%). In line with the test set results, the three methods agree on only 2.3% (*n* = 88) of cases in the entire cohort, providing more evidence of wide discrepancy among different methods.

## 4. Discussion

The literature has widely omitted HF/FO in identifying ARDS (e.g., Serpa Neto et al.^18^). Le et al.^8^ refer to this departure from the Berlin criteria as one of the limitations of their study. They posit that it would be challenging to detect HF/FO using the available data without introducing bias. We performed a sensitivity analysis to estimate the effect of excluding HF/FO in ARDS detection by executing a version of the algorithm that does not include the criterion for the 300 cases in the test set. The second row in Table 6 shows the results. Evident from the results, failing to incorporate HF/FO results in a significant drop in the accuracy of the algorithm; overall accuracy decreases by 17.0%, precision by 24.4%, and F-score by 16.1%. Recall increases because the exclusion of HF/FO will produce more positive cases, leading to a lower miss rate.

**Table 6.** The effect of different components of ARDSFlag on accuracy. Arrow/values from the second row onwards show the direction/amount of change compared to the original version.

| Method | Accuracy | Specificity | Recall | Precision | $F_1$-score |
| --- | --- | --- | --- | --- | --- |
| ARDSFlag’s baseline accuracy | 89.0% | 91.7% | 80.3% | 75.0% | 77.6% |
| ARDSFlag without the HF/FO classifier | ↓ -17.0% | ↓ -26.6% | ↑ 14.1% | ↓ -29.4% | ↓ -16.1% |
| ARDSFlag without tracheostomy as a measure of acute onset | ↓ -0.7% | ↓ -0.9% | 0.0% | ↓ -1.9% | ↓ -1.0% |
| ARDSFlag without the limit on the time of onset as a measure of acute onset | ↓ -0.7% | ↓ -1.3% | ↑ 1.4% | ↓ -2.5% | ↓ -0.7% |
| ARDSFlag without requiring a minimum 48h mechanical ventilation | ↓ -4.0% | ↓ -5.2% | 0.0% | ↓ -10.2% | ↓ -5.9% |
| ARDSFlag with a hypothetically perfect classifier for BI | ↑ 1.7% | ↑ 1.3% | ↑ 2.8% | ↑ 3.7% | ↑ 3.3% |

The third and fourth rows in Table 6 provide details on the marginal effect of acuteness measures. We used two measures to evaluate the acute onset of ARDS. First, a tracheostomy procedure within seven days of admission violates acuteness. Without this criterion, two true negative cases would be misclassified as positive. Consequently, the marginal effect of using tracheostomy as a proxy for acuity on the model’s overall accuracy is 0.7%, and on recall is 1.1%. The second criterion was the time difference between onset and the earliest time when PEEP≥ 5. As mentioned earlier, the algorithm requires this time difference to be less than seven days. By eliminating this criterion, the verdict changes for four cases; three true negative cases would move to the false positive set, and one false negative case would be resolved. Thus, this parameter has a net positive effect on the overall accuracy (89.0%-88.3%=0.7%) and the *F*_1_ score (77.6%-76.8%=0.8%).

ARDSFlag requires a minimum of 48h of mechanical ventilation unless the patient expires or opts for terminal elective extubation. By removing this condition, 12 true negative cases will be misclassified as positive, reducing the overall accuracy and *F*_1_ score by, 89.0%-85.0%=4% and 77.6%-71.7% =5.9%, respectively.

Due to the architecture of ARDSFlag, a misclassification by the BI classifier does not always lead to an ARDS misclassification. For instance, missing the evidence of BI in one radiology report may be offset by finding the evidence in another report. In the manual review of all test set cases, we found two false negatives and three false positives that were caused by BI misclassification. As shown in the last row of Table 6, the BI classifier’s imperfection has led to a 1.7% drop in the overall accuracy and a 3.3% reduction in *F*_1_ score.

Further to the above sensitivity analysis, it is worthwhile to evaluate the distribution of time of ARDS onset and its severity. Figure 4 shows the two distributions for two patient populations: the test set (*n* = 300) and the entire study cohort (*n* = 19,534). As shown in the figure, most ARDS cases arise early in patients’ clinical course in both the test set and the cohort. This correlates clinically with the usual abrupt onset of ARDS following a precipitating cause.

**Figure 4.**
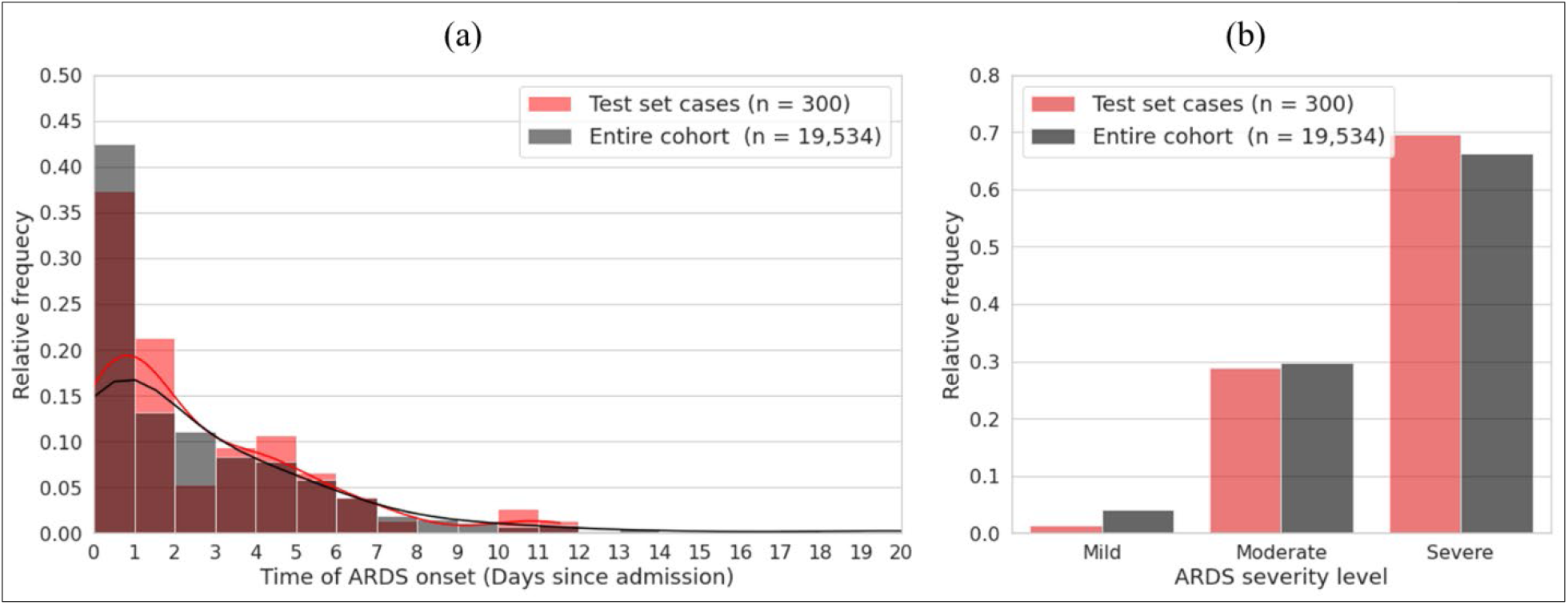
**a**. Distribution of time of ARDS onset (days since admission) for the test set and the entire study cohort. **b**. Distribution of ARDS severity in the test set and the entire study cohort.

Despite the overall accuracy and precision of the algorithm, there will be cases that go unidentified as ARDS. In one specific example reviewed, a patient with a history of pneumonectomy was deemed not to have ARDS by the algorithm. Even though the case was classified as ARDS based on clinician review, the algorithm would never identify it as such because the disease was technically unilateral. In addition, some cases showed initial evidence of cardiogenic edema. However, after aggressive diuresis, bilateral infiltrates remained present in repeat radiographic studies, and the patients were still profoundly hypoxemic. In these instances, the clinicians diagnosed the patients with ARDS. However, the algorithm would identify them as fluid overload cases and return a negative result. Regardless of the effectiveness of an algorithm, there will be nuances that will result in discrepancies between a calculated result and a clinician’s assessment.

## Data Availability

The 400 admissions that were manually labeled for ARDS are available in supplementary files. The data supporting this study's findings are available from the corresponding author upon reasonable request. The MIMIC-III database is publicly available (refer to https://mimic.mit.edu/docs/gettingstarted/ for instructions).

## 5. Limitations

A notable limitation of this type of work is that we are emulating the Berlin criteria for ARDS. By design, this prevents the inclusion of rapidity of improvement and response to diuretics over time in someone who does not have preexisting heart failure. As such, if we retrospectively analyze some ARDS-positive cases, we see patients in whom the hypoxemia resolved after a few days of diuretic therapy. Therefore, in retrospect, these cases were most likely due to pulmonary edema and not ARDS. However, prospectively, this would not have been known. Perhaps including other relevant clinical data such as fever or leukocytosis in the criteria would help exclude such cases.

A general limitation of any algorithm will be that it does not take into account nuanced details of a case that a clinician will be able to analyze. As stated, a patient who rapidly improves with diuretics would initially be classified as positive for ARDS without initial evidence of cardiac disease. This could also be true of renal patients who improve with hemodialysis. Another limitation is that the algorithm classifies bilateral infiltrates based on reports and not by actual image interpretation. There are sometimes cases where a report mentions bilateral disease, but on review of imaging, a clinician may determine that there is minimal bibasilar atelectasis. These nuances would only likely be identified by a clinician on review of a specific case, and as such is a limitation of the algorithm.

## 6. Data availability

The 400 admissions that were manually labeled for ARDS are available in supplementary files along with ARDSFlag, Serpa Neto et al., and ICD results. The data supporting this study’s findings are available from the corresponding author upon reasonable request. The MIMIC-III database is publicly available (refer to https://mimic.mit.edu/docs/gettingstarted/forinstructions).

## 7. Code availability

The data preprocessing and analysis scripts are available on GitHub and will be freely shared with academic investigators for non-commercial research.

## 8. Ethics approval

The establishment of the Medical Information Mart for Intensive Care III (MIMIC-III) was approved by the Massachusetts Institute of Technology (Cambridge, MA) and Beth Israel Deaconess Medical Center (Boston, MA), and consent was obtained for the original data collection. Therefore, the ethical approval statement and the need for informed consent were waived for this manuscript.

## 10. Author Contributions

Amir Gandomi: Literature search, Study design, Coding, Data collection, Data interpretation, Writing

Phil Wu: Study design, Data collection, Data interpretation, Writing

Daniel Clement: Study design, Literature search, Data collection, Data interpretation, Writing

Jinyan Xing: Literature search, Study design, Data collection, Data interpretation, Writing

Rachel Aviv: Literature search, Data collection, Data interpretation, Writing

Matthew Federbush: Literature search, Data collection, Data interpretation, Writing

Zhiyong Yuan: Literature search, Data collection, Data interpretation, Writing

Yajun Jing: Literature search, Data collection, Data interpretation, Writing

Guangyao Wei: Literature search, Data collection, Data interpretation, Writing

Negin Hajizadeh: Literature search, Study design, Data collection, Data interpretation, Writing

## Supplementary material

**Figure S1.**
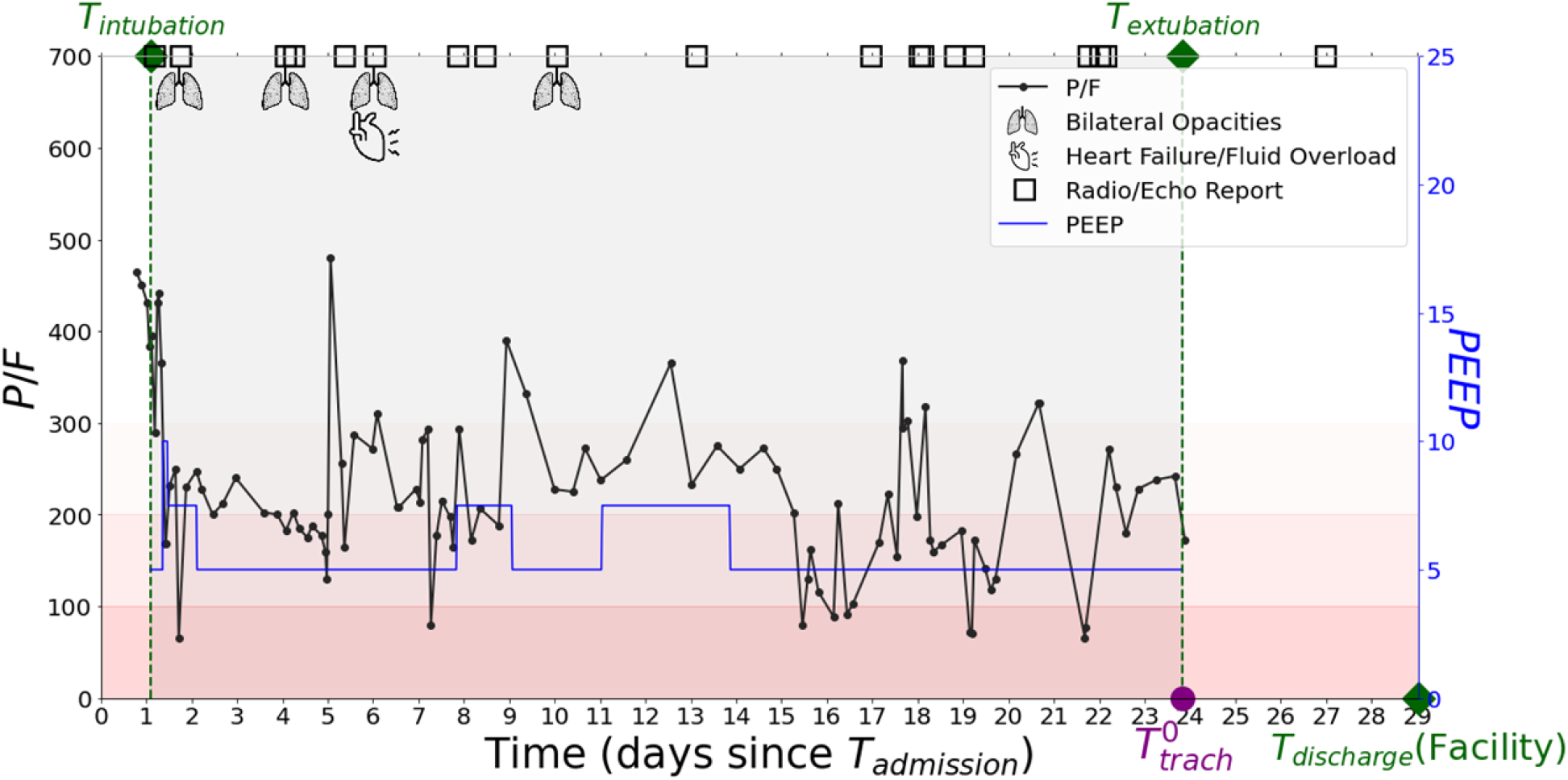
ARDS graph for HADM-ID=117986. The *ARDS graph* shows time series data on all the parameters relevant to ARDS detection, including P/F ratio, PEEP, bilateral infiltrates in the chest imaging reports, and heart failure/fluid overload in the chest imaging/echocardiogram/respiratory reports. The graph shows the earliest record of the tracheostomy procedure or oxygen delivery via tracheostomy 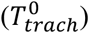. Moreover, the graph shows the times of intubation, extubation, and discharge and the discharge disposition, which is “Facility” in this example.

**Figure S2.**
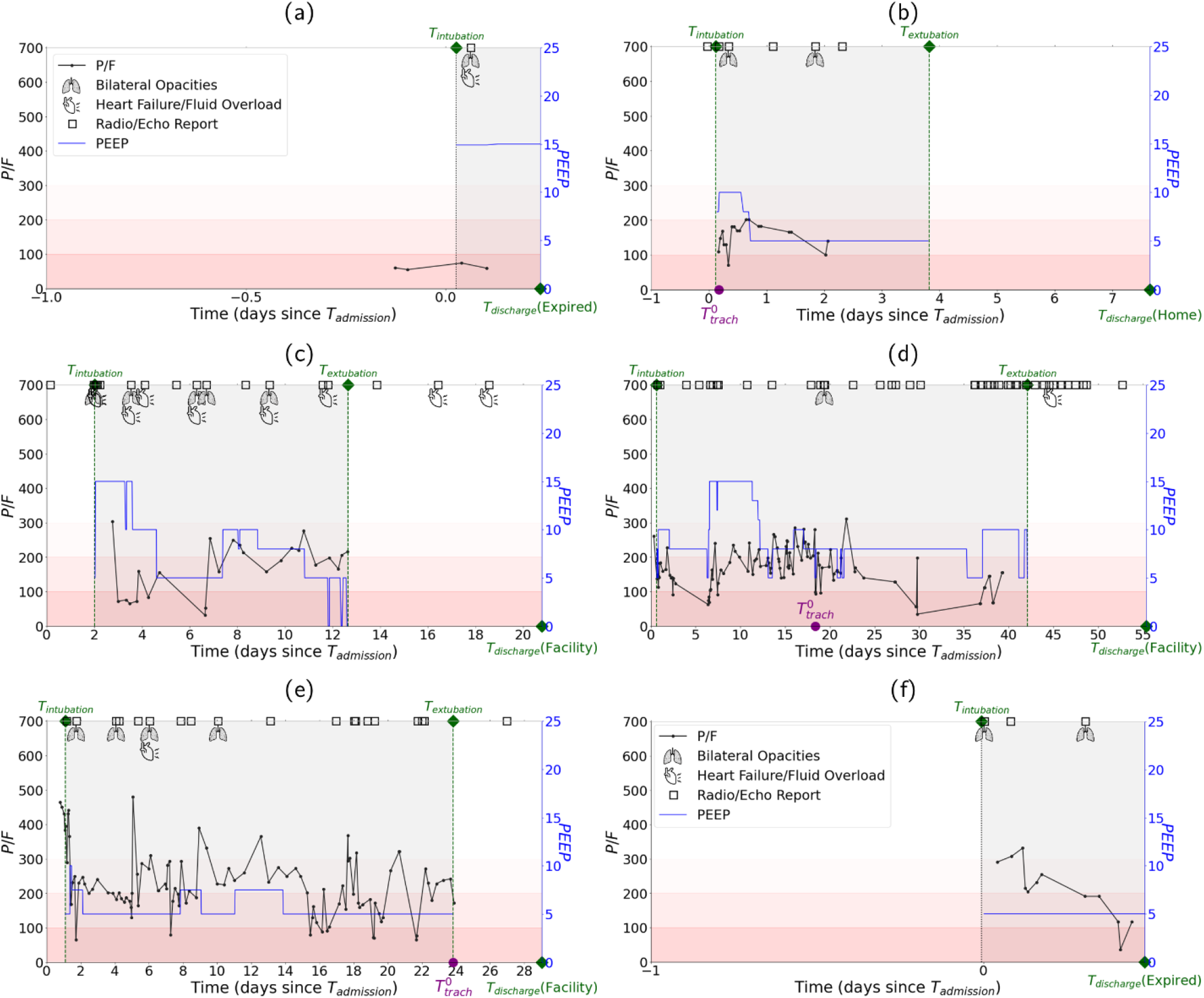
Sample cases to demonstrate the ARDS detection algorithm: **a**. HADM_ID=183612, Not ARDS because of cardiogenic cause. The short intubation length does not play a role because the patient expired within 48h of intubation. **b**. HADM_ID=105780, Not ARDS, because tracheostomy record (procedure or *O*_2_ delivery) upon admission violates the acute onset condition. **c**. HADM_ID=138174, Not ARDS because of cardiogenic cause. **d**. HADM_ID=115324, Not ARDS because the onset time (i.e., the first time P/F and PEEP condition is satisfied within bilateral infiltrates window, which is around Day 20) is beyond seven days after receiving PEEP≥ 5. **e**. HADM_ID=117986, Not ARDS because of cardiogenic cause; the first evidence of heart failure is detected around Day 6 (i.e., 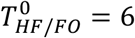, which renders heart failure as the origin of any edema observed from Day 1 onward 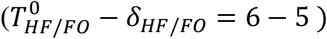. **f**. HADM_ID=126573, ARDS even though the length of intubation is less than 48 hours because the patient expires in that period.

**Figure S3.**
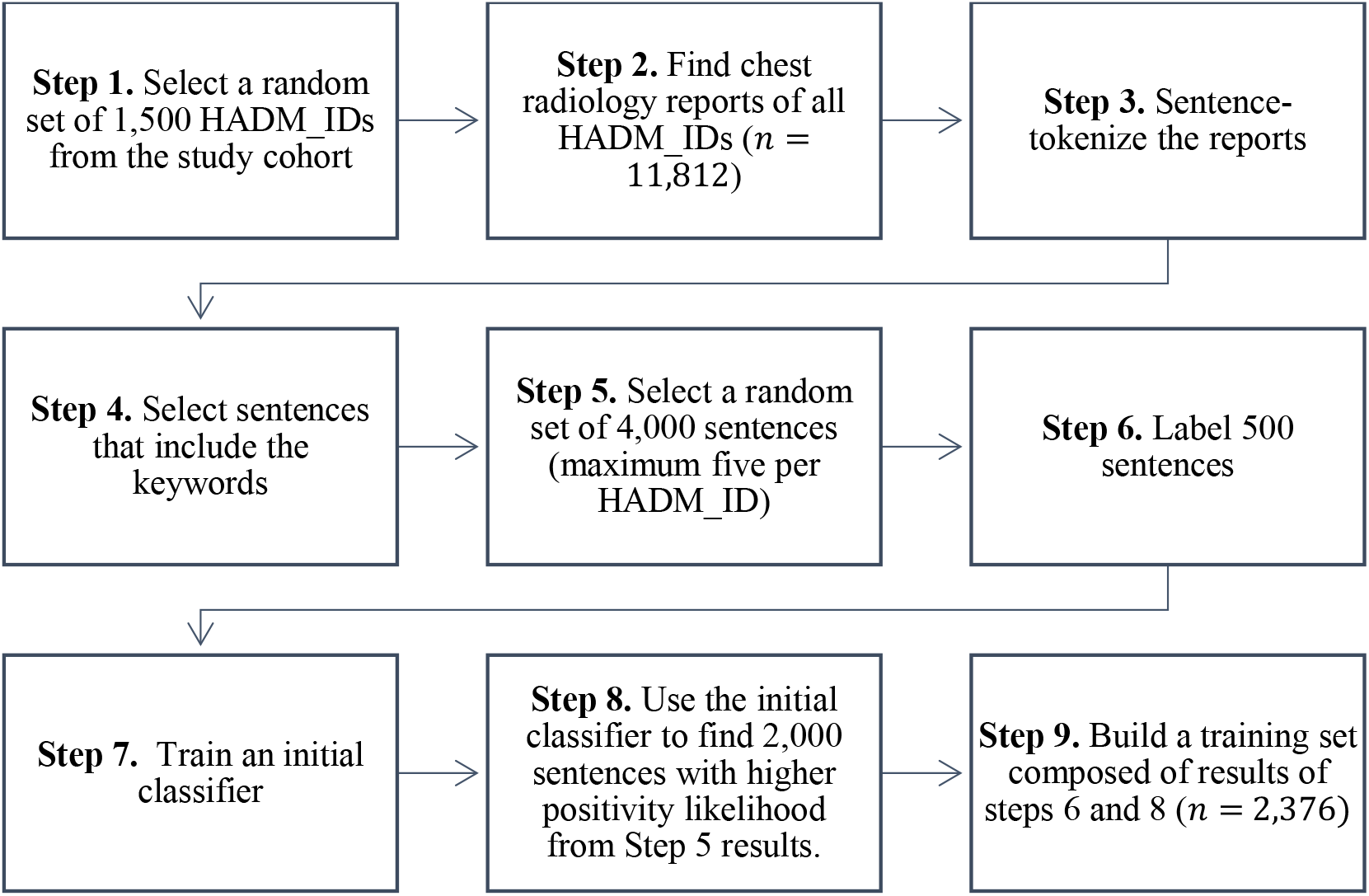
The process of developing the training set for detection of bilateral infiltrates. We used the following rule for sentence selection in Step 4: Use of any of the words in Set 1 (see below) <u>or</u> co-occurrence of at least one word from Set 2 and one from Set 3. Set 1 = {*opacity, infiltrate, consolidation, airspace disease, pneumonia, aspiration, ARDS, respiratory distress syndrome*}, Set 2 = {*bilateral, biapical, bibasilar, widespread, diffuse, perihilar, multifocal, extensive, both, lungs, left, right* }, and Set 3 = {*marking, infection, pattern, density, abnormality, haziness, hazy, process*}. Regular expressions were used to include keywords’ variations (e.g., *wide-spread, air-space disease*, and *opacities*). To create a more balanced dataset, we built an initial classifier based on 500 sentences labeled by one clinician and used it to generate 2,000 sentences that were more likely to be positive. The final training set included 2,376 sentences, 500 from Step 6 and 2,000 from Step 8, minus duplicates and non-sentences. Two clinicians labeled all sentences, resulting in 938 positive and 1,438 negative sentences.

**Figure S4.**
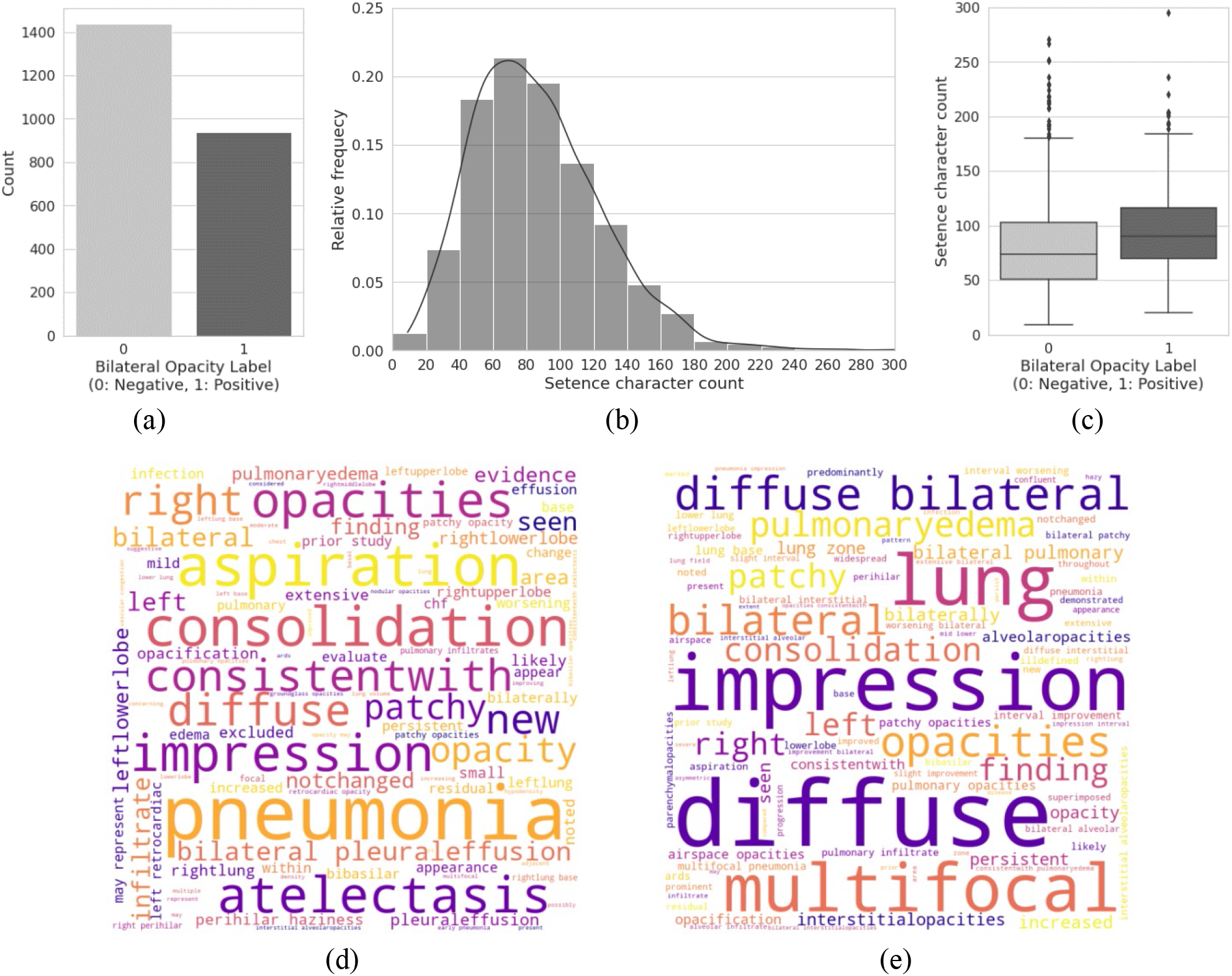
Describing the training set for bilateral infiltrates detection: **a**. frequency of positive and negative examples (*n* = 2,376); **b**. Histogram of the length of examples; **c**. distribution of length of examples by label; **d**. word cloud of unigrams and bigrams in negative examples; **e**. word cloud of unigrams and bigrams in positive examples.

**Figure S5.**
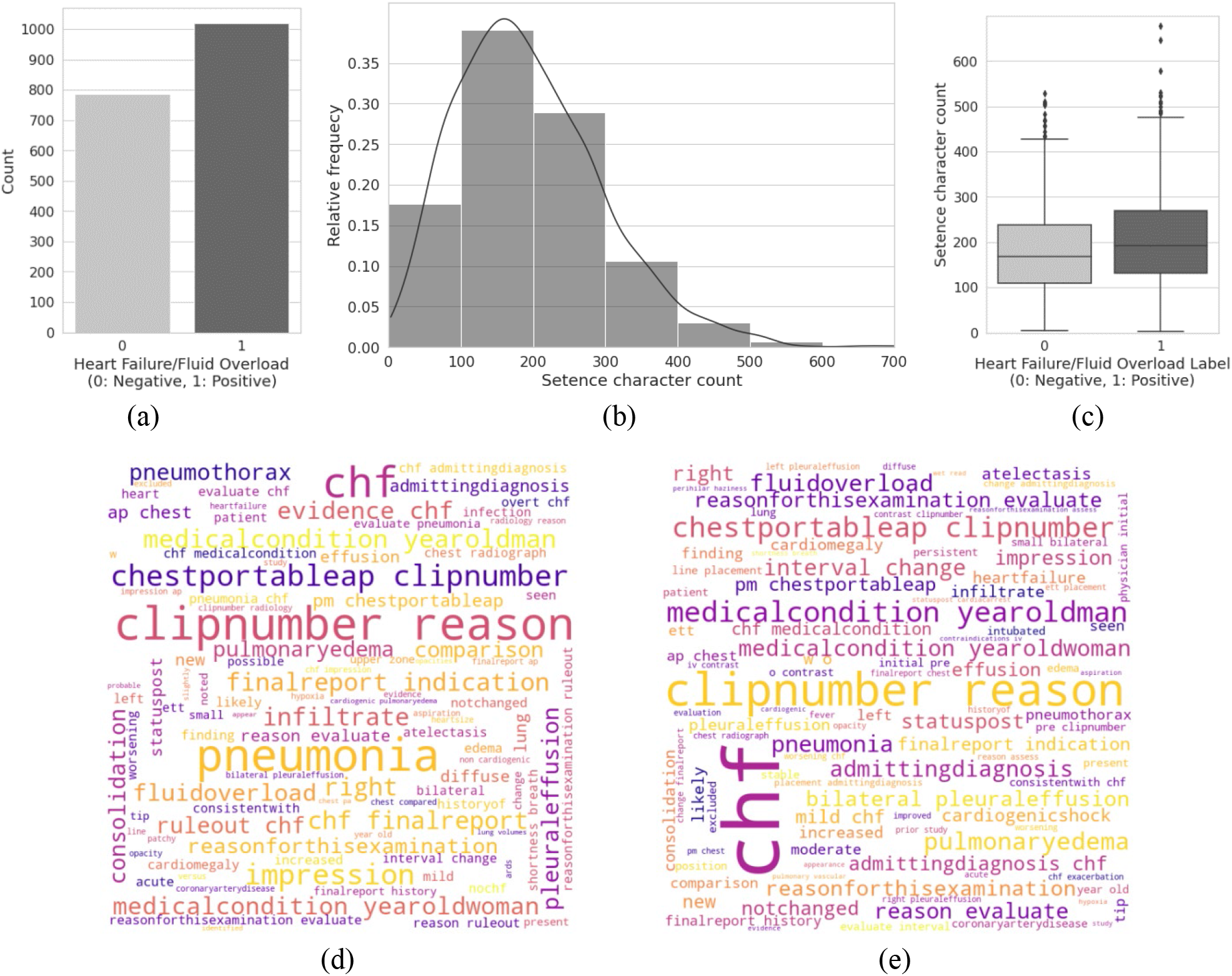
Describing the training set for heart failure/fluid overload detection: **a**. frequency of positive and negative examples (*n* = 1,808); **b**. Histogram of the length of examples; **c**. distribution of length of examples by label; **d**. word cloud of unigrams and bigrams in negative examples; **e**. Word cloud of unigrams and bigrams in positive examples.

**Table S1.** Classification pipeline hyperparameters for bilateral infiltrates detection (optimal values are highlighted)

| Step | Approach | Parameters |
| --- | --- | --- |
| Text preparation | – | - Stop words = {Standard English, **Custom list of stopwords**}<br>- Stemming = {on, **off**} |
| Vectorization | **TF-IDF** | - $n$-gram range = {only unigrams, **unigrams and bigrams**, unigrams and bigrams and trigrams}<br>- Exclude terms that occur in over $m\%$ of documents: $m = \{30, 50, 70, 90, 100\}$<br>- Exclude terms that occur in less than $n\%$ of documents: $n = \{0, 10\}$<br>- Keep only top $x$ features (ordered based on total frequency): $x = \{3000, 4000, 5000, 6000, 7000, \infty\}$<br>- Use inverse-document-frequency reweighting: **Yes**, No} |
|  | Word Embedding | - SVD dimensions: {50, 100} |
| Classification | **Stochastic gradient descent (SGD)** | - Loss function = {Hinge, Log, **Modified Huber**, Squared Hinge, Perceptron}<br>- Regularization term = **$L_2$**, Elastic net}<br>- Maximum epochs = {50, 100, **200**}<br>- Termination tolerance = {0.001, **0.0001**, 0.00001}<br>- Regularization parameter ($\alpha$) = **0.001**, 0.0001, 0.00001} |
|  | Support vector machine | - Kernel = {Linear, Polynomial, RBF}<br>- Maximum solver iterations = {10, 50, 80} |
|  | Logistic regression | - Penalty = $L_2$, Elastic net}<br>- Maximum epochs = {10, 50, 80, 250}<br>- Class weights = {balanced, {Negative:0.1, Positive:0.9}, {Negative:0.5, Positive:0.5}} |
|  | Multi-layer perceptron | - Hidden layer size = {1, 2}<br>- Number of neurons in the hidden layers = {(100), (50,50)}<br>- Activation function = {identity, logistic, tanh}<br>- Learning rate updates = {constant, adaptive}<br>- Regularization parameter ($\alpha$) = {0.00001, 0.000001} |
|  | Random forest | - Number of trees = {100, 150}<br>- Maximum depth = {None, 10, 100, 1000}<br>- Minimum samples required to split = {5, 10, 50}<br>- Minimum samples required to be at a leaf node = {1, 3, 5}<br>- Number of features to use to find the best split = {auto, sqrt, log2} |
|  | XGBoost | - Learning objective = {logistic, hinge}<br>- Feature selector = {shuffle, random, greedy} |

## References

1. Jafari, D. et al. Trajectories of hypoxemia and pulmonary mechanics of COVID-19 ARDS in the NorthCARDS dataset. BMC Pulm. Med. 22, 51 (2022).

2. Klann, J. G. et al. Validation of an internationally derived patient severity phenotype to support COVID-19 analytics from electronic health record data. J. Am. Med. Inform. Assoc. 28, 1411–1420 (2021).

3. Bernard, G. R. et al. The American-European Consensus Conference on ARDS. Definitions, Mechanisms, Relevant Outcomes, and Clinical Trial Coordination. Am. J. Respir. Crit. Care Med. 149, 818–824 (1994).

4. Thompson, B. T. & Moss, M. A New Definition for the Acute Respiratory Distress Syndrome. Semin. Respir. Crit. Care Med. 34, 441–447 (2013).

5. Beitler, J. R. et al. Personalized medicine for ARDS: The 2035 research agenda. Intensive Care Med. 42, 756–767 (2016).

6. Bellani, G. et al. Epidemiology, Patterns of Care, and Mortality for Patients With Acute Respiratory Distress Syndrome in Intensive Care Units in 50 Countries. JAMA 315, 788–800 (2016).

7. Fan, E., Brodie, D. & Slutsky, A. S. Acute Respiratory Distress Syndrome: Advances in Diagnosis and Treatment. JAMA 319, 698–710 (2018).

8. Le, S. et al. Supervised Machine Learning for the Early Prediction of Acute Respiratory Distress Syndrome (ARDS). J. Crit. Care 60, 96–102 (2020).

9. Rubenfeld, G. D., Cooper, C., Carter, G., Thompson, B. T. & Hudson, L. D. Barriers to providing lung-protective ventilation to patients with acute lung injury. Crit. Care Med. 32, 1289–1293 (2004).

10. Papazian, L. et al. Formal guidelines: management of acute respiratory distress syndrome. Ann. Intensive Care 9, 69 (2019).

11. Kalhan, R. et al. Underuse of lung protective ventilation: analysis of potential factors to explain physician behavior. Crit. Care Med. 34, 300–306 (2006).

12. Needham, D. M. et al. Timing of low tidal volume ventilation and intensive care unit mortality in acute respiratory distress syndrome. A prospective cohort study. Am. J. Respir. Crit. Care Med. 191, 177– 185 (2015).

13. Zhang, N., Gandomi, A., Wu, P., Hirsch, J. & Hajizadeh, N. An Automated Process for ARDS Detection to Facilitate the Use of Reinforcement Machine Learning. in B46. CRITICAL CARE: ALL THINGS ARDS A3517–A3517 (American Thoracic Society, 2020). doi:10.1164/ajrccm-conference.2020.201.1_MeetingAbstracts.A3517.

14. Fernandes, M. et al. Classification of the Disposition of Patients Hospitalized with COVID-19: Reading Discharge Summaries Using Natural Language Processing. JMIR Med. Inform. 9, e25457 (2021).

15. Koenig, H. C. et al. Performance of an automated electronic acute lung injury screening system in intensive care unit patients. Crit. Care Med. 39, 98–104 (2011).

16. Afshar, M. et al. Natural language processing and machine learning to identify alcohol misuse from the electronic health record in trauma patients: Development and internal validation. J. Am. Med. Inform. Assoc. 26, 254–261 (2019).

17. Oliwa, T., Furner, B., Schmitt, J., Schneider, J. & Ridgway, J. P. Development of a predictive model for retention in HIV care using natural language processing of clinical notes. J. Am. Med. Inform. Assoc. 28, 104–112 (2021).

18. Serpa Neto, A. et al. Mechanical power of ventilation is associated with mortality in critically ill patients: an analysis of patients in two observational cohorts. Intensive Care Med. 44, 1914–1922 (2018).

19. Johnson, A. E. W. et al. MIMIC-III, A Freely Accessible Critical Care Database. Sci. Data 3, 160035 (2016).

20. Makhnevich, A. et al. A Novel Method to Improve the Identification of Time of Intubation for Retrospective EHR Data Analysis During a Time of Resource Strain, the COVID-19 Pandemic. Am. J. Med. Qual. 37, 327–334 (2022).

21. The ARDS Definition Task Force*. Acute Respiratory Distress Syndrome: The Berlin Definition. JAMA 307, 2526–2533 (2012).

22. Sak, H. et al. Sequence Discriminative Distributed Training of Long Short-Term Memory Recurrent Neural Networks. in Interspeech (2014).

23. Schwager, E. et al. Utilizing machine learning to improve clinical trial design for acute respiratory distress syndrome. Npj Digit. Med. 4, 1–9 (2021).

24. Eworuke, E., Major, J. M. & Gilbert McClain, L. I. National incidence rates for Acute Respiratory Distress Syndrome (ARDS) and ARDS cause-specific factors in the United States (2006-2014). J. Crit. Care 47, 192–197 (2018).

25. Huang, B. et al. Mortality Prediction for Patients with Acute Respiratory Distress Syndrome Based on Machine Learning: A Population-Based Study. Ann. Transl. Med. 9, 794 (2021).

26. TenHoor, T., Mannino, D. M. & Moss, M. Risk factors for ARDS in the United States: analysis of the 1993 National Mortality Followback Study. Chest 119, 1179–1184 (2001).

27. Liu, J., Capurro, D., Nguyen, A. & Verspoor, K. Early prediction of diagnostic-related groups and estimation of hospital cost by processing clinical notes. Npj Digit. Med. 4, 1–8 (2021).

